# LESSONS FROM THE COVID-19 THIRD WAVE IN CANADA: THE IMPACT OF VARIANTS OF CONCERN AND SHIFTING DEMOGRAPHICS

**DOI:** 10.1101/2021.08.27.21261857

**Authors:** Finlay A. McAlister, Majid Nabipoor, Anna Chu, Douglas S. Lee, Lynora Saxinger, Jeffrey A. Bakal, on behalf of the CORONA Collaboration

## Abstract

**Importance:** With the emergence of more transmissible SARS-CoV-2 variants of concern (VOC), there is an urgent need for evidence about disease severity and the health care impacts of VOC in North America, particularly since a substantial proportion of the population have declined vaccination thus far.

**Objective:** To examine 30-day outcomes in Canadians infected with SARS-CoV-2 in the first year of the pandemic and to compare event rates in those with VOC versus wild-type infection.

**Design:** Retrospective cohort study using linked healthcare administrative datasets.

**Setting:** Alberta and Ontario, the two Canadian provinces that experienced the largest third wave in the spring of 2021.

**Participants:** All individuals with a positive SARS-CoV-2 reverse transcriptase polymerase chain reaction swab from March 1, 2020 until March 31, 2021, with genomic confirmation of VOC screen-positive tests during February and March 2021 (wave 3).

**Exposure of Interest:** VOC versus wild type SARS-CoV-2

**Main Outcomes and Measures:** All-cause hospitalizations or death within 30 days after a positive SARS-CoV-2 swab.

**Results:** Compared to the 372,741 individuals with SARS-CoV-2 infection between March 2020 and January 2021 (waves 1 and 2 in Canada), there was a shift in transmission towards younger patients in the 104,232 COVID-19 cases identified in wave 3. As a result, although third wave patients were more likely to be hospitalized (aOR 1.34 [1.29-1.39] in Ontario and aOR 1.53 [95%CI 1.41-1.65] in Alberta), they had shorter lengths of stay (median 5 vs. 7 days, p<0.001) and were less likely to die within 30 days (aOR 0.66 [0.60-0.71] in Ontario and aOR 0.74 [0.62-0.89] in Alberta). However, within the third wave, patients infected with VOC (91% Alpha) exhibited higher risks of death (aOR 1.52 [1.27-1.81] in Ontario and aOR 1.67 [1.13-2.48] in Alberta) and hospitalization (aOR 1.57 [1.47-1.69] in Ontario and aOR 1.88 [1.74-2.02] in Alberta) than those with wild-type SARS-CoV-2 infections during the same timeframe.

**Conclusions and Relevance:** On a population basis, the shift towards younger age groups as the COVID-19 pandemic has evolved translates into more hospitalizations but shorter lengths of stay and lower mortality risk than seen in the first 10 months of the pandemic in Canada. However, on an individual basis, infection with a VOC is associated with a higher risk of hospitalization or death than the original wild-type SARS-CoV-2 – this is important information to address vaccine hesitancy given the increasing frequency of VOC infections now.

---

Since December 2020, the World Health Organization has recognized 4 variants of concern (VOC) for SARS-CoV-2 as they are more transmissible.[1,2] While preliminary reports from the United Kingdom and Europe suggest VOC infections are more severe, there is a paucity of North American evidence.[3,4] The third wave of COVID-19 in Canada occurred between February and May 2021 and was driven by VOC, particularly Alpha (B.1.1.7), with Gamma (P1), Beta (B.1.351), and Delta (B.1.617) variants largely seen only in returning travellers. The Alpha variant has been shown to exhibit higher risks of mortality (approximately 64% in a UK case-control study)[5] and hospitalization (approximately 70% in the European Surveillance System data for the first 10 weeks of 2021)[6]. However, the UK study[5] relied solely on community tests, omitting as many as 70% of COVID-19 deaths occurring in patients diagnosed after hospital admission, and in the European study[6] less than 1% of SARS-CoV-2-positive specimens were sequenced for variants. A pre-print from Denmark also reported higher hospitalization rates with the Alpha variant, but was based on only 128 (of 1235 total) hospitalizations and the difference was only detected in adjusted analyses.[7] Thus, questions remain, and to our knowledge nothing has yet been published on disease severity or trajectories (ie. timing of hospitalizations or death) with VOC compared to the original SARS-CoV-2 clade in North America, crucial information for health system planners.

To address this gap in the literature, we designed this study to examine 30-day outcomes in Canadians infected with SARS-CoV-2 in the first 13 months of the pandemic and to compare event rates in those with VOC versus wild-type infection.

## METHODS

### Subjects and Setting

We conducted a retrospective cohort study in two of Canada’s most populous provinces - Alberta and Ontario. Canadian healthcare is a government funded single-payer system with free universal access to hospital, emergency department (ED), laboratory, and physician services and each province is the legal custodian of the health data for its citizens. We linked SARS-CoV-2 reverse transcriptase Polymerase Chain Reaction (RT PCR) testing data from the Alberta and Ontario provincial laboratories with administrative health databases in each province which capture all ED visits, hospitalizations, ICU admissions, and deaths.

### Ethics

Ethical approval for this study was granted by the University of Alberta Health Ethics Research Board (Pro00101096), with waiver of individual patient signed informed consent for Alberta data as we analyzed de-identified healthcare administrative data within the secure environment of the Alberta Strategy for Patient Oriented Research Support Unit. The use of Ontario data is authorized under section 45 of Ontario’s Personal Health Information Protection Act (PHIPA) and does not require review by another Research Ethics Board as it was analyzed within the secure environment of the Institute for Clinical Evaluative Sciences (ICES) which is a prescribed entity (ie. health information custodian) under the PHIPA. As a prescribed entity, ICES undergoes tri-annual review and approval of its privacy and security policies, procedures and practices by Ontario’s Information and Privacy Commissioner. In addition, each research project that is conducted at ICES (including this one) is also subject to internal ethical review by the ICES Privacy and Compliance Office.

### Definition of Cases, Index Date, and Outcomes

The study population included all individuals (outpatients and inpatients) with a positive SARS-CoV-2 reverse transcription polymerase chain reaction (RT-PCR) nasopharyngeal swab from March 1, 2020 until March 31, 2021, with genomic confirmation of all VOC screen-positive tests after February 7, 2021. For patients tested multiple times during our study, we only examined the data related to their first positive SARS-CoV-2 test. Index date was the date of the first positive RT-PCR test and outcomes examined included ED visits, hospitalizations, ICU admissions, and/or deaths in the first 30 days after the positive RT-PCR test.

### Covariates

We identified comorbidities for each patient (and generated their Charlson Comorbidity Scores) using standardized ICD-9 and ICD-10-CA case definitions (previously validated in Alberta and Ontario) based on all hospitalizations in the 2 years prior to and including the index date for each individual.

### Statistical Analyses

We present summary statistics stratified according to the timing of the positive SARS-CoV-2 swab and, for those detected after February 1, 2021 whether they had VOC or wild-type SARS-CoV-2 detected. We compared outcome risks after adjusting for age, sex, and Charlson Comorbidity Score (which includes the most important of the QCOVID risk score factors [https://qcovid.org/] such as diabetes, pulmonary disease, kidney disease, heart failure, neurologic disease, and cancer. All analyses were conducted using SAS 9.4 [Cary, NC, USA] within each province separately.

### Data Disclosure

To comply with each province’s Health Information Protection Act, the dataset used for this study cannot be made publicly available but requests to access the dataset from qualified researchers trained in human subject confidentiality protocols may be sent to the corresponding author (Dr. Finlay McAlister).

## RESULTS

Compared to the 372,741 individuals with SARS-CoV-2 positive samples between March 2020 and January 2021 (the Canadian first and second waves), there was a leftward shift in age distribution for the 104,232 COVID-19 cases identified in the third wave in Alberta and Ontario (Table). The percentage of test samples positive for SARS-CoV-2 was similar in all 3 waves (5.2% overall). Hospitalization rates within 30 days were higher in the third wave (5.5% vs. 5.3%, p=0.008), a difference which persisted after adjusting for the differences in demographics and comorbidity burdens (aOR 1.53 [95%CI 1.41-1.65] in Alberta and aOR 1.34 [1.29-1.39] in Ontario for third wave patients vs. earlier waves). However, hospital lengths of stay were shorter in the third wave (median 5 vs. 7 days in Alberta and 6 vs. 7 days in Ontario, both p<0.001) than in the first and second waves. Patients with COVID-19 during the third wave were also less likely to die within 30 days (0.8% vs. 2.2%, p<0.0001) than those in the first and second waves, even after adjusting for their younger age and lower comorbidity burdens (aOR 0.74 [0.62-0.89] in Alberta and aOR 0.66 [0.60-0.71] in Ontario).

**Table:**
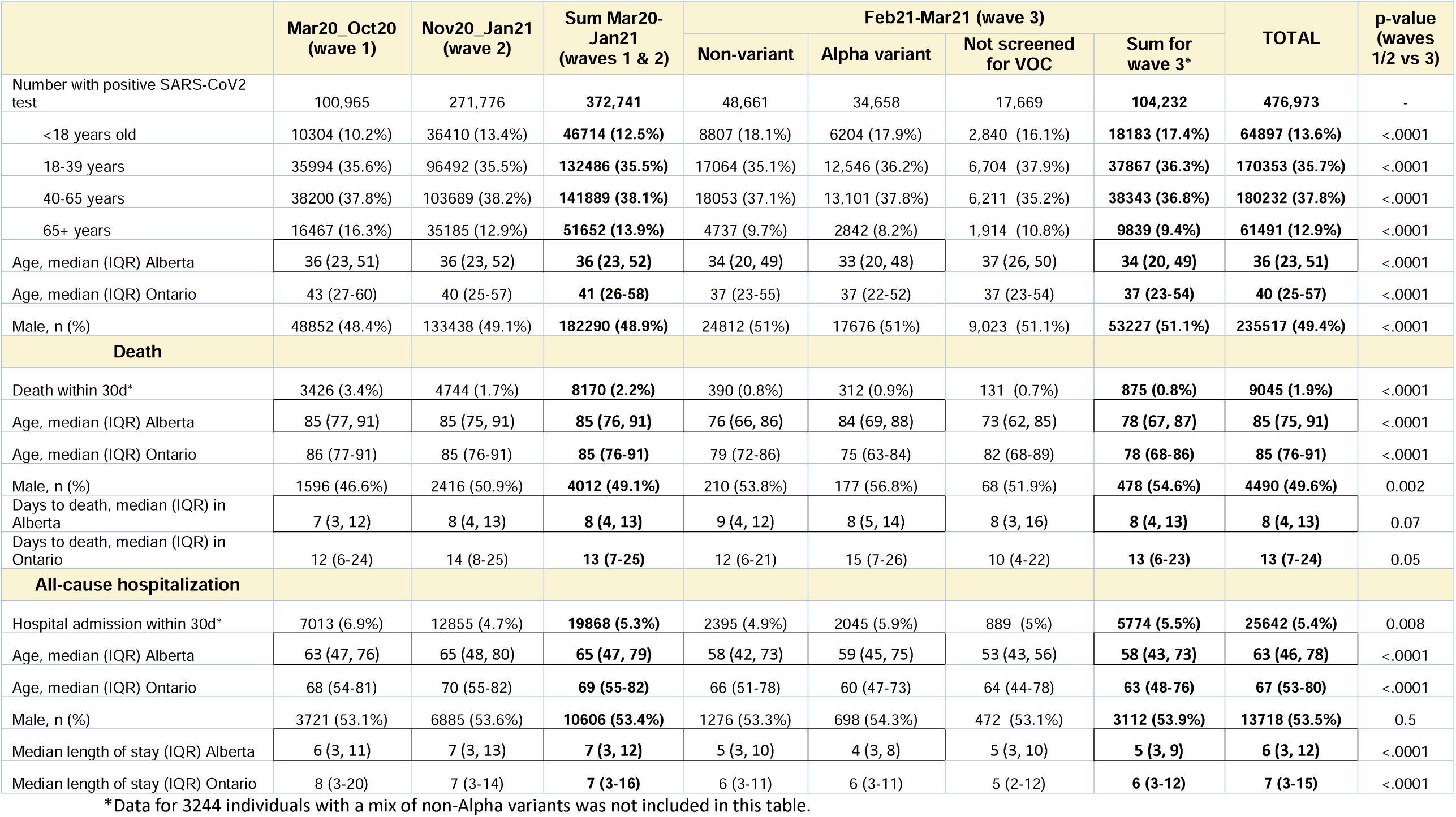
Baseline characteristics and outcomes for individuals with SARS-CoV2 infection from March 1 2020 to March 31 2021 in Alberta and Ontario, Canada.

Examining Canadian third wave data only, of the 104,232 patients with SARS-CoV-2 positive samples in February or March 2021, 86,563 (83%) were screened for VOC, of which 37902 (43.8%) were confirmed VOC positive, and 34,658 (91.4%) were the Alpha variant - only 27 delta variants were isolated. After adjusting for age, sex and comorbidities, VOC-infected patients exhibited higher 30-day risks of death (aOR 1.67 [1.13-2.48] in Alberta and aOR 1.52 [1.27-1.81] in Ontario) and hospitalization (aOR 1.57 [1.47-1.69] in Ontario and aOR 1.88 [1.74-2.02] in Alberta) than those with non-VOC infections in the same timeframe. However, lengths of hospital stays and times between positive swab and death were similar among patients with VOC versus wild-type infections in the third wave.

## DISCUSSION

Our data provides one of the first descriptions of disease severity phenotypes for SARS-CoV-2 VOC in North America, which is important for public health messaging and for health system planners. Consistent with effect estimates from the UK and Europe,[4,6] we found a 52% to 67% higher mortality risk with predominantly Alpha VOC infections in Canada. However, given that the third wave in Canada affected younger patients more than the first two waves (at least partially due to vaccination eligibility criteria in early 2021), we also found that the overall COVID-19 case fatality rate was 30% lower in the third wave compared to the earlier waves, even though hospitalization risk increased by over 40%. Our finding that individuals infected with a VOC were more likely to require hospitalization than those with wild type SARS-CoV-2 is consistent with reports from Europe of a higher hospitalization risk with the VOC (over 80% of which were Alpha in those studies).[6,7] However, since time to event trajectories were similar for variant and wild type SARS-CoV-2 infections and lengths of stay were shorter, health system projections on timing of hospitalizations based on incident case counts should not be affected by the emergence of VOC.

A major strength of our study is that the majority of SARS-CoV-2-positive specimens during the Alberta and Ontario third wave were screened for VOC with genomic confirmation, compared with very low proportions in other studies describing VOC disease severity (for example, only 0.7% in the recent report from the European Surveillance System[6]). A limitation of our analysis is that it is largely based on Alpha variant infections and additional research is needed to determine whether other VOC exert similar risk.[3,6,7] In particular, given emerging evidence that the Delta variant replicates faster and Delta-infected individuals have much higher viral loads (over 1,200 fold in a recent preprint)[8], there is an urgent need to define the phenotype of Delta variant disease. As with other studies comparing VOC and non-VOC infections, our sampling frame may result in overestimates of absolute risks since minimally symptomatic patients are less likely to be tested. However, we examined all positive community cases, which is less biased than studying only hospitalized cases (which prior studies have done) and sample positivity rates were similar in all 3 waves (approximately 5%). Unfortunately, the duration of the pre-symptomatic stage (and the frequency of asymptomatic cases) with different SARS-CoV-2 clades are not yet described and cannot be assessed using our dataset. Finally, although we do not have data on vaccination status in our dataset, initial vaccine roll-out in Canada in January and February 2021 focused on long term care residents, the very elderly, indigenous adults, and front line health care workers only and it was not until mid-March that vaccination eligibility criteria expanded to include other groups in both provinces.[9,10]

While genomic monitoring for the evolution of SARS-CoV-2 VOCs is crucial,[11] we believe it is equally important to describe disease expression and outcomes among patients infected with VOC to fully understand both the disease phenotype and the anticipated burdens for the healthcare system. Although preliminary data regarding the effectiveness of current vaccines against hospitalization and death from the VOC are encouraging,[12–14] continued assessment of disease severity phenotypes in different jurisdictions in vaccinated and unvaccinated individuals and as VOCs evolve is important.

For health system planners, the shift in COVID-19 disease epidemiology towards younger age groups as the pandemic evolves translates into more hospitalizations but shorter lengths of stay and lower mortality risk than seen in the first 10 months of the pandemic. However, demonstration that the VOC are associated with substantially higher risk of hospitalization or death than infections with the original wild-type strain in North America is important for individual patient counselling and public information campaigns reinforcing social distancing guidelines and addressing vaccine hesitancy.

## Supporting information

STROBE checklist

## Data Availability

To comply with each province Health Information Protection Act the dataset used for this study cannot be made publicly available but requests to access the dataset from qualified researchers trained in human subject confidentiality protocols may be sent to the corresponding author.

## Acknowledgements

The Ontario portion of this study was supported by and conducted within ICES, which is funded by an annual grant from the Ontario Ministry of Health (MOH) and the Ministry of Long-Term Care (MLTC). Parts of this material are based on data and information provided by the Canadian Institute for Health Information (CIHI) and MOH. These data were provided to ICES under section 45 of PHIPA and may only be used for the “purpose of analysis or compiling statistical information with respect to the management of, evaluation or monitoring of, the allocation of resources to or planning for all or part of the health system. The analyses, opinions, results, and conclusions expressed herein are solely those of the authors and do not necessarily reflect those of the funding or data sources; no endorsement is intended or should be inferred; no endorsement is intended or should be inferred. The Alberta portion of this study was supported by and conducted within the Alberta Strategy for Patient Oriented Research Support Unit. This study is based in part on data provided by Alberta Health and Alberta Health Services. The interpretation and conclusions contained herein are those of the researchers and do not represent the views of the Government of Alberta or Alberta Health Services. Neither the Government of Alberta nor Alberta Health Services express any opinion in relation to this study.

## REFERENCES

1. Fontanet A, Autran B, Lina B, Kieny MP, Karim SSA, Sridhar D. SARS-CoV-2 variants and ending the COVID-19 pandemic. Lancet 2021;397:952–954. doi: 10.1016/S0140-6736(21)00370-6.

2. CDC. Emerging SARS-CoV-2 Variants. 2021; published online Jan 29. https://www.cdc.gov/coronavirus/2019-ncov/more/science-and-research/scientific-brief-emerging-variants.html (Accessed June 29, 2021).

3. Curran J, Dol J, Boulos L, Somerville M, McCulloch H. Public Health and Health Systems Impacts of SARS-CoV-2 Variants of Concern: A Rapid Scoping Review [last updated June 6, 2021]. medRxiv 2021.05.20.21257517; doi: https://doi.org/10.1101/2021.05.20.21257517

4. Davies NG, Jarvis CI, CMMID COVID-19 Working Group, et al. Increased mortality in community-tested cases of SARS-CoV-2 lineage B.1.1.7. Nature 2021;March 15: https://doi.org/10.1038/s41586-021-03426-1

5. Challen R, Brooks-Pollock E, Read J M, Dyson L, Tsaneva-Atanasova K, Danon L et al. Risk of mortality in patients infected with SARS-CoV2 variant of concern 202012/1: matched cohort study. BMJ 2021;372:n579

6. Funk T, Pharris A, Spiteri G, Bundle N, Melidou A, Carr M, et al. Characteristics of SARS-CoV-2 variants of concern B.1.1.7, B.1.351 or P.1: data from seven EU/EEA countries, weeks 38/2020 to 10/2021. Euro Surveillance 2021;26:pii=2100348. https://doi.org/10.2807/1560-7917.ES.2021.26.16.2100348

7. Bager P, Wohlfahrt J, Fonager J, et al. Increased risk of hospitalisation associated with infection with SARS-CoV-2 lineage B.1.1.7 in Denmark. Lancet Preprint at https://doi.org/10.2139/ssrn.3792894 (Accessed June 29, 2021).

8. Li B, Deng A, Li K, Hu Y, Li Z, Xiong Q, et al. Viral infection and transmission in a large, well-traced outbreak caused by the SARS-CoV-2 Delta variant. medRxiv 2021.07.07.21260122; doi: https://doi.org/10.1101/2021.07.07.21260122

9. Ismail SJ, Zhao L, Tunis MC, et al. Key populations for early COVID-19 immunization: preliminary guidance for policy. CMAJ 2020;192:E1620–32.

10. McAlister FA, Bushnik T, Leung AA, Saxinger L. Informing COVID-19 vaccination priorities based on the prevalence of risk factors among adults in Canada. CMAJ. 2021;193:E617–E621.

11. Alam I, Radovanovic A, Incitti R, Kamau AA, Alarawi M, Azhar EI, et al. CovMT: an interactive SARS-CoV-2 mutation tracker, with a focus on critical variants. Lancet Infect Dis 2021. [PMID: 33571446] doi: 10.1016/S1473-3099(21)00078-5

12. Sah P, Vilches TN, Moghadas SM, Fitzpatrick MC, Sing BH, Hotez PJ, Galvani AP. Accelerated vaccine rollout is imperative to mitigate highly transmissible COVID-19 variants. E Clinical Medicine 2021;35:100865

13. Iorio A, Little J, Linkins L, Abdelkader W, Bennett D, Lavis JN. COVID-19 living evidence synthesis #6 (version 6.14): What is the efficacy and effectiveness of available COVID-19 vaccines in general and specifically for variants of concern? Hamilton: Health Information Research Unit, July 28 2021.

14. Lopez Bernal J, Andrews N, Gower C, Gallagher E, Simmons R, Thelwall S, Stowe J, Tessier E, Groves N, Dabrera G, Myers R, Campbell CNJ, Amirthalingam G, Edmunds M, Zambon M, Brown KE, Hopkins S, Chand M, Ramsay M. Effectiveness of Covid-19 Vaccines against the B.1.617.2 (Delta) Variant. N Engl J Med. 2021 Jul 21:NEJMoa2108891. doi: 10.1056/NEJMoa2108891. Epub ahead of print. PMID: 34289274; PMCID: PMC8314739.

